# Deep Learning Prediction of Biomarkers from Echocardiogram Videos

**DOI:** 10.1101/2021.02.03.21251080

**Authors:** J Weston Hughes, Neal Yuan, Bryan He, Jiahong Ouyang, Joseph Ebinger, Patrick Botting, Jasper Lee, John Theurer, James E. Tooley, Koen Neiman, Matthew P. Lungren, David Liang, Ingela Schnittger, Bob Harrington, Jonathan H. Chen, Euan A. Ashley, Susan Cheng, David Ouyang, James Y. Zou

**Author notes:** Co-senior author (David Ouyang, James Y. Zou).

## Abstract

Laboratory blood testing is routinely used to assay biomarkers to provide information on physiologic state beyond what clinicians can evaluate from interpreting medical imaging. We hypothesized that deep learning interpretation of echocardiogram videos can provide additional value in understanding disease states and can predict common biomarkers results. Using 70,066 echocardiograms and associated biomarker results from 39,460 patients, we developed EchoNet-Labs, a video-based deep learning algorithm to predict anemia, elevated B-type natriuretic peptide (BNP), troponin I, and blood urea nitrogen (BUN), and abnormal levels in ten additional lab tests. On held-out test data across different healthcare systems, EchoNet-Labs achieved an area under the curve (AUC) of 0.80 in predicting anemia, 0.82 in predicting elevated BNP, 0.75 in predicting elevated troponin I, and 0.69 in predicting elevated BUN. We further demonstrate the utility of the model in predicting abnormalities in 10 additional lab tests. We investigate the features necessary for EchoNet-Labs to make successful predictions and identify potential prediction mechanisms for each biomarker using well-known and novel explainability techniques. These results show that deep learning applied to diagnostic imaging can provide additional clinical value and identify phenotypic information beyond current imaging interpretation methods.

## Introduction

Diagnostic medical testing provides insight into human physiology and disease conditions, with testing ranging from blood based biomarkers and genetics testing to imaging studies that provide deep insight into anatomy and changes over time^1–3^. Blood based laboratory testing is a fundamental tool for disease diagnosis and management as changes in assayable biomarkers can be some of the earliest signs of physiological perturbations^5–7^. Despite the frequent utilization of both laboratory testing and medical imaging in routine clinical practice, the deeper connections between medical images and biomarkers values are relatively underexplored^4^. It remains unknown whether routinely obtained imaging studies might contain information that can broadly predict common biomarker values and more deeply inform clinicians about the patient condition.

Recent advances in Artificial Intelligence have shown that deep learning applied to medical images can identify phenotypes beyond what is currently possible by observation from human clinicians alone ^8–11^. Such discoveries have spanned across a variety of imaging modalities in many medical specialties and have uncovered imaging correlates for a wide range of disease states, molecular signatures, and physiologic conditions ^12–15^. Given that orthogonal and complimentary information is obtained from the many different forms of diagnostic testing, subtle associations and relationships can be missed in conventional clinical assessment.

Echocardiograms, or cardiac ultrasounds, are the most common form of cardiovascular imaging, combining rapid image acquisition, lack of ionizing radiation, and high temporal resolution to capture spatiotemporal information on cardiac motion and function^16,17^. Previous works have shown deep learning based assessment of echocardiograms can identify physiological state and hints of both systemic as well as cardiac diseases ^9,18,19^. In the extremes, abnormal blood chemistry can influence cardiac function^20^, and over time, structure, but it is unknown whether transient or subtle variations biomarkers are reflected in the physiologic state that can be extracted from medical imaging. A deep learning assessment of frequently obtained, no radiation, low cost, and information dense imaging, such as echocardiogram videos, could provide additional diagnostic information that alleviates the need for other invasive, costly, or burdensome forms of testing. This is the first demonstration that echocardiograms can be used to detect abnormal blood biomarkers through deep learning analysis of the ultrasound videos, and our artificial intelligence algorithms generalize imaging and biomarker results across healthcare systems.

## Results

### Data Curation

We curated a dataset of 70,066 echocardiogram videos from 39,460 patients at Stanford Medicine and 1,300 videos from 819 patients from Cedars-Sinai Medical Center. The echocardiogram videos were matched with 14 associated blood-based biomarker tests. Both biomarker assays with particular relevance to cardiac function and myocyte damage, such as B-type Natriuretic Peptide (BNP) and troponin I, as well as biomarkers of systemic physiology such as hemoglobin and blood urea nitrogen (BUN), were paired with echocardiogram videos for model training. Echocardiogram videos from Stanford Medicine were preprocessed and curated for apical 4-chamber view videos and divided based on patient into 59,434 training, 5,319 validation, and 5,313 internal test examples. An additional dataset of 1,301 apical 4-chamber view videos from Cedars-Sinai Medical Center were never seen during model training and served as an hold-out external test set for this study. The data is described in Supplemental Tables 1 and 2.

### Video-based deep learning model to predict biomarkers

We developed a deep learning framework, EchoNet-Labs, to answer whether medical imaging might be able to predict biomarker values and whether these results generalize across different clinical settings and healthcare systems (Figure 1). EchoNet-Labs is a convolutional neural network with residual connections and spatiotemporal convolutions that provides a beat-by-beat estimate for biomarker values. Extending our prior work on deep learning applied to echocardiogram videos^19^, EchoNet-Labs incorporates both spatial and temporal information to perform both regression and classification tasks.

**Figure 1:**
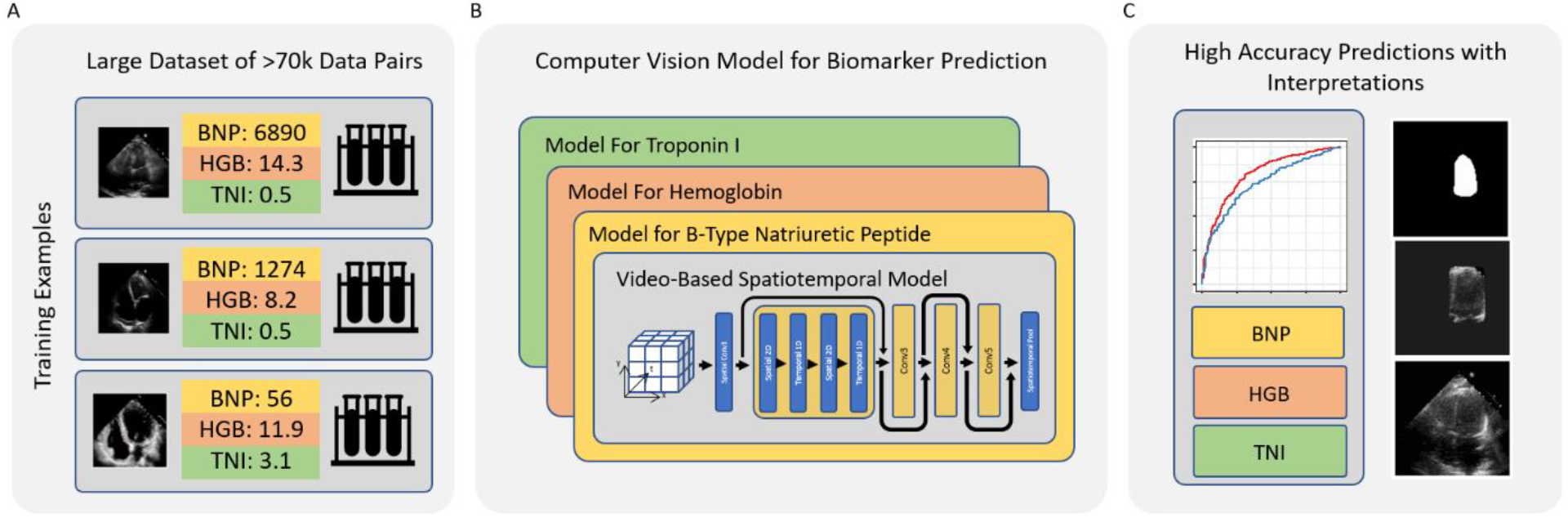
Overview of EchoNet-Labs system and study design. A. A training dataset of over seventy thousand echocardiogram videos and paired biomarker values from the same patient were used to train a video-based AI system for prediction of laboratory values. B. Our deep learning based AI system used spatio-temporal convolutions to infer biomarker values from both anatomic (spatial) and physiologic (temporal) information contained with echocardiogram videos. C. To understand the relative importance of spatial and temporal information, ablation datasets removing texture, motion, and extracardiac structures were adopted to perform interpretations experiments.

### Evaluation of model performance

On the held-out test set of patients from Stanford Medicine that was not previously seen during model training, EchoNet-Labs predicted biomarker values from echocardiogram videos with high sensitivity and specificity (Figure 2). EchoNet-Labs achieved an area under the curve (AUC) of 0.80 (0.79-0.81) in predicting anemia (low hemoglobin), of 0.86 (0.85-0.88) in predicting elevated BNP, of 0.75 (0.73-0.78) in predicting elevated troponin I, of 0.74 (0.72-0.76) in predicting elevated BUN, and up to 0.72 in predicting abnormalities in ten other common laboratory tests (Supplementary Table 3). To provide context for these results, we also trained a model to predict each biomarker using demographics and standard quantitative metrics from echocardiograms, such as left ventricular ejection fraction. This baseline model achieved AUC of 0.60, 0.72, 0.69, and 0.62 for predicting anemia, BNP, troponin I, and BUN respectively, which is substantially lower than EchoNet-Labs’ performance. This comparison suggests that EchoNet-Labs captures novel features in the videos beyond correlates of patient demographics and commonly annotated cardiac features.

**Figure 2:**
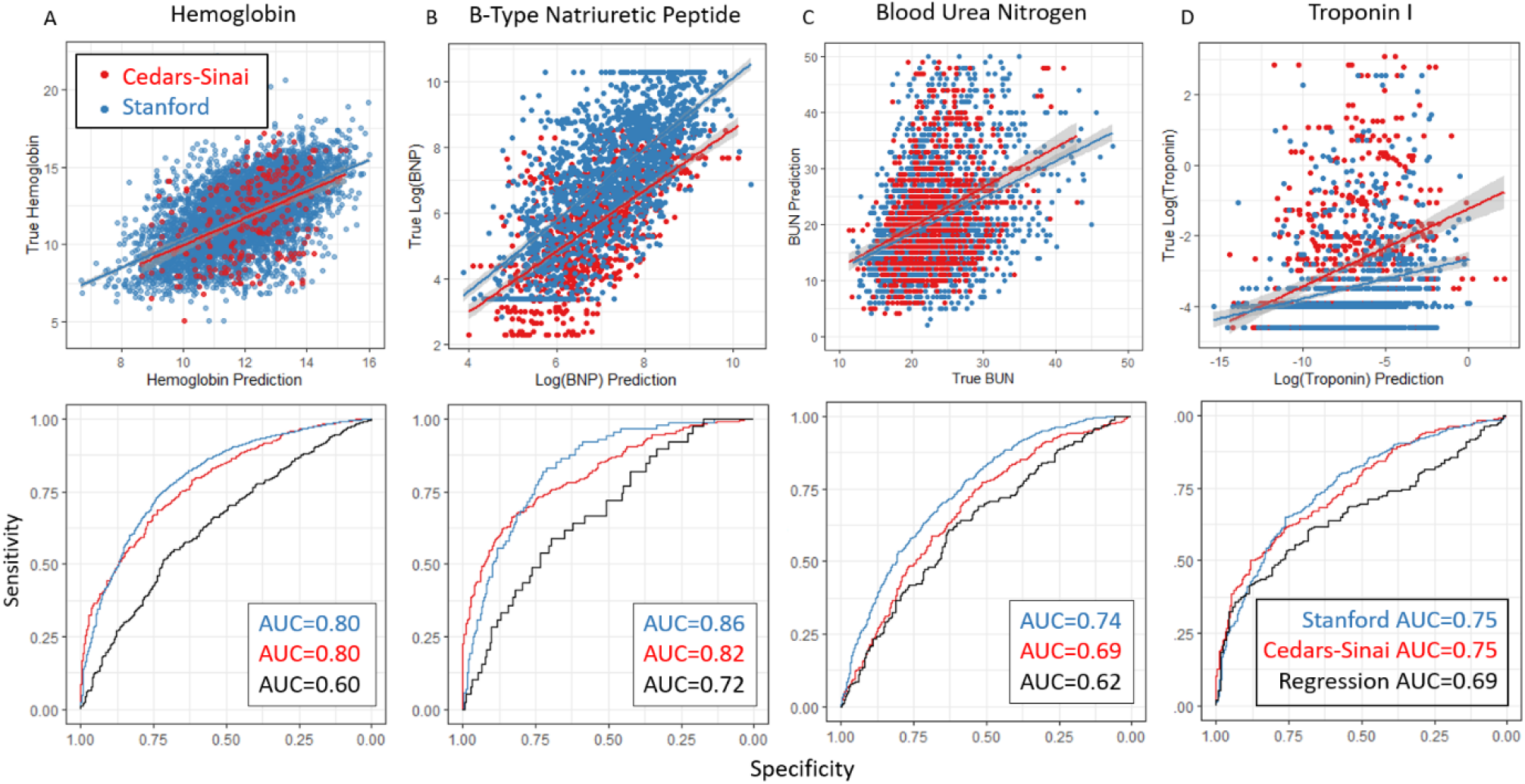
Performance of EchoNet-Labs on Internal and External Test Datasets. A-D. Scatterplots (top) and receiver-operating characteristic (ROC) curves (bottom) for prediction of (A) hemoglobin, (B) B-Type Natriuretic Peptide, (C) Blood Urea Nitrogen, and (D) Troponin I. Blue points and curves denote to a held-out test set of patients from Stanford Medicine not previously seen during model training. Red points and curves denote to performance on the external test set from Cedars-Sinai Medical Center. Black curves denote a benchmark with linear regression using demographics and echocardiogram features (LVEF, RVSP, Heart Rate) on the Stanford test set.

### External testing on cross-health system data

To assess the cross-healthcare-system reliability of the model, EchoNet-Labs was additionally tested, without any tuning, on an external test dataset of 1,301 patients from Cedars-Sinai Medical Center. On the external test dataset, EchoNet-Labs achieved an AUC of 0.80 (0.77-0.82) in predicting anemia, of 0.82 (0.79-0.84) in predicting elevated BNP, of 0.75 (0.72-0.78) in predicting elevated troponin I, and of 0.69 (0.66-0.71) in predicting elevated BUN, which is similar to the model’s accuracy on the Stanford test patients. This analysis further supports the generalizability of EchoNet-Labs across different settings.

### Analyzing model performance and understanding high importance imaging features

To clarify which features are most relevant to EchoNet-Labs’ prediction of each biomarker, we trained a series of models on various transformations of the input data to remove different types of information (Figure 3). EchoNet-Labs achieved high performance in predicting elevated BNP, Troponin I, and BUN based on video of only the region around the left ventricle (AUCs of 0.88, 0.74, and 0.74 respectively, compared to 0.89, 0.73, and 0.75 on the full video), suggesting information from that region alone might be sufficient for biomarker prediction. EchoNet-Labs’ performance was slightly worse but still quite accurate when making predictions based on video of only the tracing of the left ventricle endothelium (AUCs of 0.84, 0.71, and 0.71), and based on a single randomly selected frame of video (AUCs of 0.84, 0.69, and 0.71). This demonstrates that both the motion of the ventricle in the absence of fine-grained pixel and texture, and the fine-grained pixel and texture in the absence of motion information, each contain a large amount of the information. When evaluating results of predicting anemia, model performance depended more on texture information as performance was greatly limited by restricting input to the left ventricular border (AUC dropped from 0.81 to 0.67, versus 0.73 on single frame). Finally, we performed sensitivity analysis with regard to training sample size (Supp. Figure 1). Even with large sample sizes of up to 58,000 training examples, we do not see an inflection in improvement in performance, suggesting EchoNet-Labs can be improved with additional training examples.

**Figure 3:**
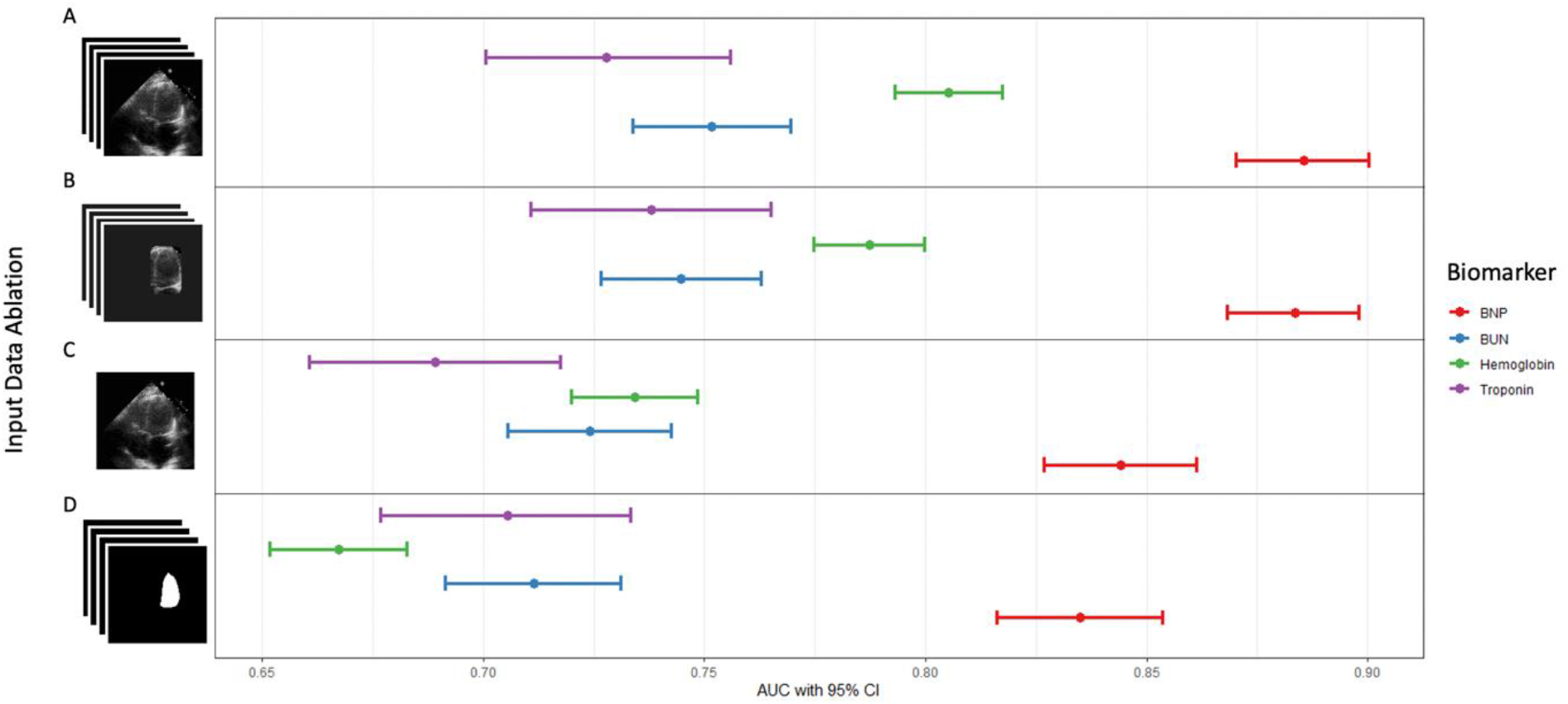
EchoNet-Labs input ablations and impact on model performance. Experiments showing performance of models trained on ablated input data that hides specific information. (A) Results on standard video input. (B) Results with input where region outside of the left ventricle are obscured. (C) Results with removing temporal information with single frame input. (D) Results with removing texture information and showing location of the left ventricle. For each ablation setting, a separate model was trained on that type of ablated data to quantify the information content in the data. The width of each bar indicates the bootstrap 95% CI for each prediction AUC.

## Discussion

EchoNet-Labs is a video-based deep learning algorithm that achieves state-of-the-art prediction of biomarkers from echocardiogram videos. Using 70,066 echocardiogram videos and paired biomarker results, EchoNet-Labs has high accuracy in predicting abnormal hemoglobin, BNP, troponin I, and BUN, and this performance was superior to a model using traditional risk factors. The model performance was robust to changing the clinical environment, and experiments degrading the input data show EchoNet-Labs incorporates both motion and texture based information for its assessment. The results of this study support a growing body of literature highlighting that deep learning analysis of medical imaging can identify correlative findings of systemic physiology that was previously thought to be only obtained from orthogonal diagnostic testing^8,23^.

With deep learning models and model interpretation techniques, our study highlights the association between imaging phenotypes and biomarkers of both cardiovascular and systemic disease. Echocardiogram videos are commonly used to diagnose heart failure, which has a strong association with some biomarkers (e.g. BNP) and can help explain the strong performance of EchoNet-Labs for BNP. Similarly troponin I is most abundantly found in cardiac myocardium and is frequently used as a marker of myocardial injury and myocardial infarction.

Surprisingly, we show disease states and biomarkers not directly related to cardiovascular function can be readily predicted from echocardiogram videos, extending prior work in other modalities that show medical imaging might have additional value in understanding the patient condition^21,22^. Other biomarkers, such as hemoglobin and BUN, are associated with systemic disease but now shown convincingly to be predicted accurately both by imaging and electrical signals of the cardiovascular system^23^. How hemoglobin and BUN values are associated with cardiac motion has not been previously characterized. Our findings provide the first evidence that variation in these values are visually detectable in heart motion. The physiological response to anemia includes tachycardia and compensatory changes in cardiac function which could be picked up by deep learning models in the prediction of abnormal hemoglobin. Improved understanding of the close relationships between imaging and laboratory testing can lead to further understanding of the relationship between imaging phenotypes and disease processes.

Performance in deep learning prediction of biomarkers varied considerably by biomarker, with the highest AUCs for some biomarkers associated with cardiovascular disease (troponin I and BNP) while other blood chemistries had less dynamic range and were not able to be predicted confidently. Integrating echocardiograms and lab values can help inform the interpretation of both tests and in doing so provide an overall more accurate picture of disease. It may also help clarify how certain lab abnormalities might correspond to changes in cardiac structure and function. Additionally, our experiments suggest EchoNet-Labs can continue to be improved with additional training examples, which suggest a promising direction of further exploration.

If proven to be reliable, laboratory prediction from fast, cheap imaging could be useful in numerous clinical contexts. In the emergency room, point-of-care echocardiography is already used to triage procedures and assist medical decision-making in medical emergencies. While laboratory testing requires phlebotomy and processing, often offsite, ultrasound is often readily available and rapidly attained, even in resource-limited settings. As a rapid adjunct to conventional testing, EchoNet-Labs can help stratify patients by risk or guide medical decision making in obtaining expensive laboratory testing when there is low clinical suspicion and low probability for abnormal testing results.

## Methods

### Data curation details

From the population of patients who received at least one lab test and one echocardiogram at Stanford in the last 20 years, we randomly selected 70,066 echocardiogram studies. A single apical-4-chamber 2D gray-scale video was identified from each study and used to represent the study for mapping to laboratory values. Previously described methods ^19^ were used to preprocess echocardiogram videos to standard resolution and remove extra information outside of the ultrasound sector such as text, ECG and respirometer data, as well as identifying information. Laboratory values were extracted from the electronic health record and paired with the representative echocardiogram video. The 70,066 videos were split by patient identifier into 59,434 videos for training, 5319 videos for validation, and 5313 videos for internal testing, such that the same patient never appeared in multiple splits of the data. During training, if there are multiple videos from the same patient, we treat them as individual samples for training. An additional external test dataset of 1,301 videos with corresponding biomarker results was obtained from Cedars-Sinai Medical Center and processed using the same pipeline without further fine tuning. Binary thresholds for model performance assessment were determined by the reference range of the particular laboratory’s assay, and for biomarkers with significant variance (BNP, Troponin I, CRP, ALT, AST), model training was performed on the logarithm of the result value. During model training, the lab closest in time to each video was used as the training label, and videos were excluded from training if the patient did not have a corresponding video-laboratory value pair. In the validation and test sets, the same process was applied with the additional constraint that only labels acquired within 30 days of the echocardiogram were included. In the case of CRP, a window of 365 days was used to increase sample size. This research was approved by the Stanford University and Cedars-Sinai Medical Center Institutional Review Boards.

### Model development and training

Models were built using Python 3.8 and PyTorch 1.4. Extending on previous work^19^, EchoNet-Labs uses a (2+1)D-ResNet consisting of 34 layers of alternating spatial and temporal convolutions in a ResNet structure^24^. We chose the same hyperparameter configuration as in previous work^19^ and found that architecture choice (e.g. R3D and MC3) and temporal step size (e.g. 1/1, 1/2, or 1/4 the sampling rate of the original video) do not significantly affect results. All models were pretrained on the Kinetics-400 dataset^25^. Independent video regression models were trained for each lab value, taking as input a randomly selected 32×112×112 sub-video and predicting the lab value. We also explored training a single model to predict all values through multi-task learning, but found for key lab values that training individual models performed better. Log values were predicted for BNP, CRP, ALT, and AST due to skewed distributions. Abnormal values were defined using reference thresholds for the particular assay in the calculating area under receiver operating characteristic (AUC) curve. Notably, the EchoNet-Labs prediction for BNP was trained on paired echocardiogram videos and NT-proBNP results from Stanford Medicine, and tested on BNP data from Cedars-Sinai, which uses a different assay. Videos were augmented during training by randomly shifting by up to 12 pixels.

The models were trained to minimize the mean squared error between the prediction and true lab value. Model training used a stochastic gradient descent optimizer with an initial learning rate of 0.001, momentum of 0.9, and batch size of 20 for 45 epochs. The learning rate was decayed by a factor of 0.1 every 15 epochs. Prediction was set up as a binary classification task, predicting normal versus abnormal lab value, based on standard thresholds. For these biomarkers, clinicians recognize inherent heterogeneity on retesting and often make clinical decisions on whether broadly these biomarkers are either normal or abnormal. To understand model generalization, each model was evaluated on a held-out test set not used in any way during model development, from a set of patients completely disjoint from those used during training. Finally, for the four most successful biomarkers we report results on the Cedars-Sinai external validation dataset. For each lab, we report the AUC on the validation and test sets, with bootstrapped 95% confidence intervals.

For all models, the weights from the epoch with the lowest validation AUC was selected for final testing. Our final model averaged predictions across the entire echocardiogram video over all possible 32 frame sub-videos rather than randomly selecting one to account for potential variance between beats. We report area under the receiver operator characteristic curve (AUC) as the primary performance metric in figure 2 and supplementary figure 3. All confidence intervals are 95% confidence intervals generated by bootstrapping on the relevant test set. Predicting a single lab value with EchoNet-Labs, with all test-time augmentation, takes less than 5 seconds.

### Video content transformation

To further understand the features needed to make classifications, we retrained models for anemia, BNP, troponin I, and BUN on differently ablated inputs. For each transformation, we trained and tested on identically ablated data. To understand if motion-based features are necessary for classification, we trained and tested a model on a single randomly selected frame of each echo, repeated 32 times in a video to fairly compare to other 3D resnet models. To understand if the motion of the left ventricle on its own is sufficient for classification, we trained and tested a model on a video of the segmented outline of the left ventricle generated by Echonet-Dynamic, with none of the original video data present. To understand if only the information in and around the left ventricle is sufficient to classify, we trained and tested a model with all data outside of a bounding box around the left ventricle obscured. To produce a video of just the left ventricle, we found the smallest bounding box which contained the left ventricle in all frames, expanded it by 5 pixels, and set all pixels outside of that region to 0.

### Comparison to benchmark model

One way a model might learn to predict a biomarker value would be to use covariates which are known to be contained in echocardiogram data, and use those covariates as well as discrete demographic information to predict the biomarker value. Age and sex have been previously shown to be predictable from echocardiogram videos with high accuaracy^9,19^, and echocardiogram videos contain information about left ventricular ejection fraction, heart rate, and right ventricular systolic pressure. To determine if the model truly learned novel features, we trained a linear regression model using these demographics and echocardiography derived metrics to compare with EchoNet-Labs.

### Effect of Dataset Size

To understand the impact of input sample size on EchoNet-Labs, we trained separate models with datasets at different sized subsets for each biomarker. Models were trained by randomly selecting 1000, 2000, 4000, and 8000 training examples for each model. Upward trends were observed for all values as dataset size increased without a clear inflection point, suggesting that further growth in the dataset size could further increase accuracy. In particular, doubling the size of the datasets consistently leads to uniform increases, suggesting that partnering with other healthcare systems to produce multiplicatively larger datasets would lead to further gains in accuracy.

## Data Availability

All of the code for EchoNet-Labs will be available at https://github.com/echonet/ after publication. Matched laboratory values will be provided through our publicly available EchoNet dataset at https://echonet.github.io/.

## Supplementary Information

**Supplementary Table 1.** Lab statistics. Summary statistics for each dataset and biomarker. The number of patients and mean (standard deviation) of the laboratory value is provided for the train, validation, Stanford test and the external Cedars-Sinai test cohorts. *Model was trained on the logarithm of the value given high variance of the biomarker values.

|  | Train |  | Validation |  | Stanford Test |  | Cedars-Sinai Test |  |
| --- | --- | --- | --- | --- | --- | --- | --- | --- |
|  | n | mean (sd) | n | mean (sd) | n | mean (sd) | n | mean (sd) |
| BNP* | 30434 | 3490 (6537) | 1614 | 4143 (6712) | 1667 | 4387 (7108) | 898 | 740 (1078) |
| Hemoglobin | 58116 | 12.4 (2.2) | 4932 | 12.0 (2.3) | 4911 | 12.0 (2.3) | 1226 | 11.5 (2.4) |
| Troponin* | 23298 | 0.21 (1.88) | 988 | 0.29 (2.01) | 1033 | 0.20 (1.19) | 683 | 0.57 (2.30) |
| BUN | 44958 | 22 (14) | 2503 | 24 (16) | 2570 | 23 (16) | 1056 | 28 (20) |
| Creatinine | 58884 | 1.26 (1.18) | 5199 | 1.29 (1.18) | 5192 | 1.30 (1.19) |  |  |
| ALT* | 56964 | 48 (156) | 4666 | 53 (197) | 4706 | 53 (224) |  |  |
| CRP* | 15876 | 8.44 (22.50) | 983 | 8.07 (21.85) | 870 | 10.08 (24.73) |  |  |
| AST* | 56883 | 46 (315) | 4653 | 57 (402) | 4685 | 57 (430) |  |  |
| Sodium | 58722 | 138 (3.66) | 5169 | 138 (4.03) | 5157 | 137 (3.90) |  |  |
| WBC | 57973 | 7.76 (8.56) | 4924 | 8.49 (12.74) | 4903 | 8.25 (9.77) |  |  |
| A1c | 38247 | 6.00 (1.27) | 1676 | 6.34 (1.58) | 1748 | 6.34 (2.93) |  |  |
| AlkPhos | 34762 | 142 (109) | 1876 | 174 (146) | 1897 | 179 (148) |  |  |
| Platelet | 58003 | 214 (93) | 4922 | 214 (105) | 4902 | 210 (106) |  |  |
| Chloride | 58700 | 103 (4.13) | 5144 | 103 (4.45) | 5143 | 102 (4.40) |  |  |

**Supplementary Table 2.** Lab cutoffs and number of patients above cutoff. Each lab value was binarized to be above or below the given threshold for the prediction purpose. For purposes of classification of abnormal results, biomarkers were classified based on the reference range of assay.

| Biomarker | Threshold | Train<br>Number (Proportion)<br>above threshold | Validation<br>Number (Proportion)<br>above threshold | Test<br>Number (Proportion)<br>above threshold |
| --- | --- | --- | --- | --- |
| BNP | 300 | 21011 (0.69) | 1179 (0.73) | 1261 (0.76) |
| Hemoglobin | 10 | 50256 (0.86) | 3912 (0.79) | 3880 (0.79) |
| Troponin | 0.0001 | 14599 (0.63) | 629 (0.64) | 619 (0.60) |
| BUN | 30 | 8088 (0.18) | 542 (0.22) | 504 (0.20) |
| Creatinine | 1.5 | 10212 (0.17) | 991 (0.19) | 1018 (0.20) |
| ALT | 200 | 781 (0.01) | 91 (0.02) | 87 (0.02) |
| CRP | 2 | 8612 (0.54) | 545 (0.55) | 527 (0.61) |
| AST | 200 | 779 (0.01) | 93 (0.02) | 98 (0.02) |
| Sodium | 130 | 58366 (0.99) | 5003 (0.97) | 4983 (0.97) |
| WBC | 12 | 5114 (0.09) | 583 (0.12) | 569 (0.12) |
| A1c | 6.5 | 8004 (0.21) | 539 (0.32) | 540 (0.31) |
| AlkPhos | 200 | 3991 (0.11) | 359 (0.19) | 426 (0.22) |
| Platelet | 150 | 47180 (0.81) | 3772 (0.77) | 3655 (0.75) |
| Chloride | 105 | 18169 (0.31) | 1617 (0.31) | 1532 (0.30) |

**Supplementary Table 3:** EchoNet-Labs performance on internal and external test data. Area under ROC curve for each lab value on the internal Stanford test set and the external Cedars-Sinai test sets. *CRP results are based on a 365 day window to increase sample size.

|  | Stanford Test AUC |  | Cedars-Sinai Test AUC |  |
| --- | --- | --- | --- | --- |
| BNP | 0.86 | (0.85-0.88) | 0.82 | (0.79-0.84) |
| Hemoglobin | 0.8 | (0.79-0.81) | 0.8 | (0.77-0.82) |
| Troponin | 0.75 | (0.73-0.78) | 0.75 | (0.72-0.78) |
| BUN | 0.74 | (0.72-0.76) | 0.69 | (0.66-0.71) |
| Cr | 0.72 | (0.70-0.73) |  |  |
| ALT | 0.72 | (0.67-0.76) |  |  |
| CRP* | 0.69 | (0.66-0.72) |  |  |
| AST | 0.69 | (0.65-0.74) |  |  |
| Sodium | 0.67 | (0.64-0.70) |  |  |
| WBC | 0.64 | (0.62-0.66) |  |  |
| A1c | 0.63 | (0.61-0.66) |  |  |
| AlkPhos | 0.62 | (0.59-0.65) |  |  |
| Platelet | 0.61 | (0.60-0.63) |  |  |
| Chloride | 0.54 | (0.53-0.56) |  |  |

**Supplementary Figure 1:**
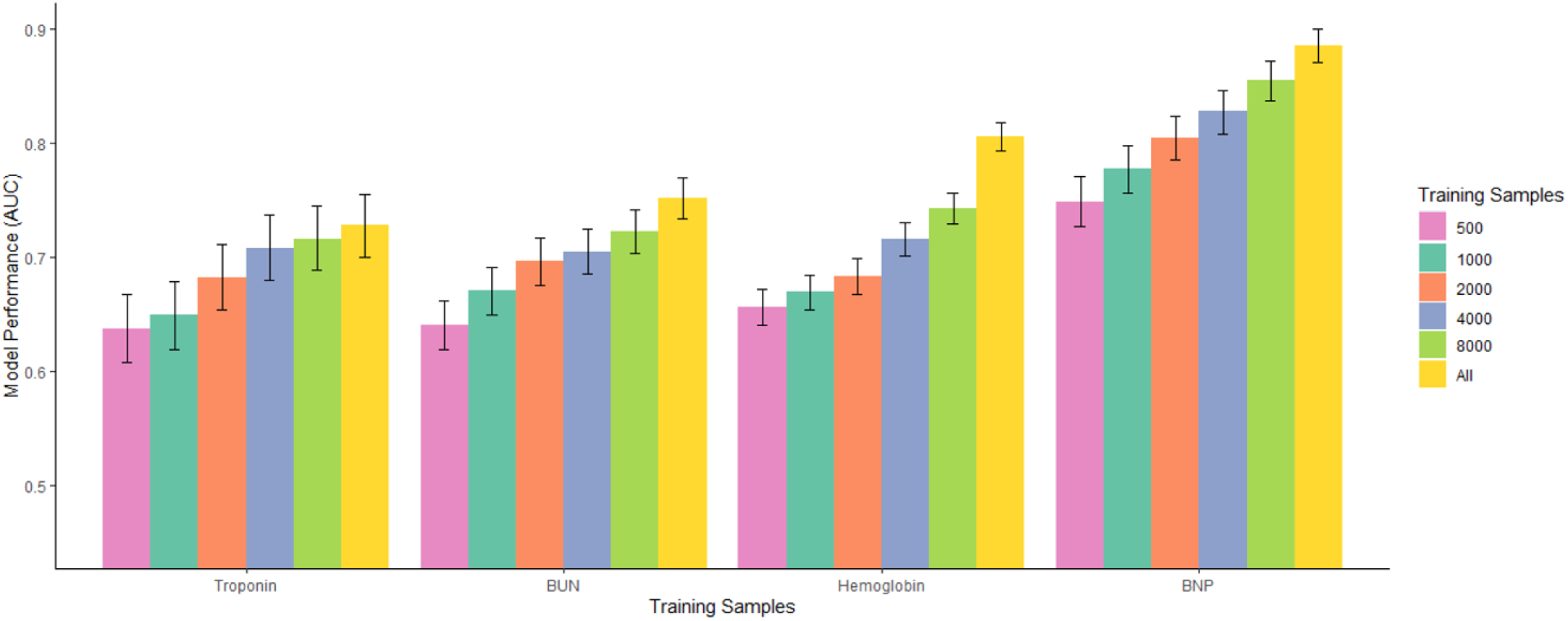
Performance of EchoNet-Labs when varying training dataset size. Training samples refers to the number of unique video-biomarker pairs used during training, where All represents the maximum sample size for the entire curated dataset.

